# What Do Child Abuse and Neglect Medical Evaluation Consultation Notes Tell Us?

**DOI:** 10.1101/2022.06.22.22276783

**Authors:** Megan Golonka, Yuerong Liu, Rosie Rohrs, Jennie Copeland, Jessalyn Byrd, Laura Stilwell, Carter Crew, Molly Kuehn, Elizabeth Snyder-Fickler, Jillian Hurst, Kelly Evans, Lindsay Terrell, Elizabeth Gifford

## Abstract

Child abuse and neglect (CAN) medical experts provide specialized multidisciplinary care to children when there is concern for maltreatment. Their clinical notes contain valuable information on child- and family-level factors, clinical concerns, and service placements that may inform the needed supports for the family. We created and implemented a coding system for data abstraction from these notes. Participants were 1,397 children ages 0-17 years referred for a consultation with a CAN medical provider at an urban teaching and research hospital between March 2013 and December 2017. Coding themes were developed using an interdisciplinary team-based approach to qualitative analysis, and descriptive results are presented using a developmental-contextual framework. This study demonstrates the potential value of developing a coding system to assess characteristics and patterns from CAN medical provider notes, which could be helpful in improving quality of care and prevention and detection of child abuse.

## Introduction

Child maltreatment is a common, serious, and costly problem in the United States. One in seven children experiences physical abuse, sexual abuse, emotional abuse, and/or neglect each year, resulting in 1,840 fatalities in 2019 (Centers for Disease Control and Prevention, 2021). In addition to suffering immediate physical, emotional, and/or psychological injuries, children who experience maltreatment face long-term consequences that impact their health and well-being. Child maltreatment is more likely to occur when a family’s stressors and risks outweigh their supports and protective factors (Belsky, 1993). Examining these factors in context is crucial. A developmental-contextual approach to the ecology of child maltreatment considers the interaction of multiple factors at multiple contextual levels (e.g., developmental, demographic, immediate-situational) that may increase the likelihood of child abuse and neglect (Belsky, 1993; Bronfenbrenner, 1979). Approaching the problem of child maltreatment with a developmental-contextual ecological approach and an understanding of the individual- and family-level risk and protective factors involved allows medical providers a broader view of families’ experiences. It also enables them to engage with other services in a collaborative, multi-system response.

Given the potential serious long-term effects, children who are suspected to have been maltreated require specialized, multidisciplinary care that takes contextual factors into consideration. In their efforts to perform medical evaluations for concerns of child maltreatment, child abuse and neglect (CAN) medical providers elicit social history and risk factors in addition to past medical and family history and presenting issues. Evaluations usually include obtaining histories from the child’s caregiver/s who report on social drivers of health, family and caregiver networks, family medical history, and risk factors such as interpersonal violence, criminal justice system involvement, and substance use. Additional histories may also be provided by social workers and law enforcement if they are involved. During outpatient visits, the child also undergoes a diagnostic interview. Child abuse and neglect (CAN) medical providers perform medical record reviews and physical examinations for children referred for abuse and neglect concerns, including laboratory tests, radiographic tests, and photo documentation as needed. They also provide referrals to other pediatric subspecialties such as mental health, community support, and prevention programs. CAN medical providers participate in multidisciplinary teams which may include community agencies investigating and managing abuse and neglect cases. They may also be asked to provide expert testimony (Block & Palusci, 2006). Additionally, CAN medical clinics develop and coordinate care plans for children and their families to mitigate factors that can contribute to poor outcomes.

Increasingly, this information from medical evaluations is being captured within electronic health records (EHR). These clinical notes are a rich source of information on child- and family-level factors and social drivers that may directly contribute to both the incidence of maltreatment and treatment outcomes. Such information could be extremely helpful in improving quality of care and prevention and detection of child abuse. However, given the intended purpose of these notes, it can be difficult to derive discrete data elements in a consistent and reliable manner.

Limited research has thoroughly examined the notes documented in CAN medical clinics. One such effort examined inpatient medical notes from children younger than five years who had sustained one of three specific injury types that could be suspicious for physical abuse (traumatic brain injury, long bone fracture, and skull fracture) (Olson et. al., 2018). Through qualitative content analysis, the researchers found rich social histories contained in the 730 sets of inpatient medical consultation notes provided by 32 board-certified pediatricians from 23 child abuse programs in the United States. This study provides an example of the utility of qualitatively coding CAN medical provider notes, but it is limited in the scope of developmental stages and types of maltreatment it examined.

In the current study, we expanded Olson et al.’s (2018) work by creating a coding system for abstracting and examining all aspects of comprehensive medical evaluation notes. We included children of all ages, both inpatient and outpatient records, and all types of suspected maltreatment including physical, sexual, emotional, and medical abuse, as well as neglect. We evaluated over 1,400 notes involving children and adolescents 0-17 years of age who were assessed at a hospital-based CAN medical clinic. The information in the notes was gathered through interviews by the care team using a structured template, but the notes themselves were stored without any structure in place to allow for data to be easily extracted. Here, we provide a detailed description of our protocol and considerations for developing and implementing a coding system designed to assess case characteristics, such as training and supervising coders to obtain the optimal data. We present baseline descriptive data to demonstrate the utility of variables extracted from both structured EHR data and clinical notes. Finally, we provide recommendations for further research using these methods.

## Methods

### Participants

The sample used in this study includes children ages 0-17 years who were referred for an inpatient or outpatient consultation with a CAN medical provider between March 13, 2013, and December 1, 2017. Because children are likely to receive care within the county in which they live, we limited our sample to children who resided in the same county as the hospital at the time of the visit.

### Data

Data for this study were derived from the EHR of an urban teaching and research hospital in the Southeastern United States utilizing the hospital’s secure protected analytic workspace in which sensitive EHR data can be stored and analyzed. This hospital has a CAN medical evaluation clinic for patients who are suspected to have experienced child abuse and/or neglect. We use the term “child abuse and neglect (CAN) medical providers” because our team includes child abuse pediatricians and other advanced practice providers such as nurse practitioners specializing in the care of children who have experienced maltreatment (Herold et al., 2018), as well as social workers employed by this clinic and/or Child Protective Services (CPS). The university health system institutional review board (IRB) approved this study for research purposes. We also obtained a signed research collaboration agreement from CPS that states that the data are approved for use under the oversight of the university health system IRB.

#### Medical Notes

Medical notes were recorded by expert CAN medical providers during consultations at the time of the child’s medical evaluation. Notes were generated during clinic office visits and through hospital consults. When available, the notes included information that previously existed in patients’ EHR prior to the medical evaluation for maltreatment, such as previous diagnoses and hospital stays. The sources of additional information in the notes varied by case and could include any combination of responses regarding family medical and social history gathered through a semi-structured narrative interview guided by a clinic template. Questions asked during the medical evaluation included evidence-based developmental-contextual factors that have been identified as common contributors to child maltreatment. Reporters could include any combination of caregivers, social workers, and law enforcement. In addition, each clinic visit includes a diagnostic interview for any referred focal child who is over the age of three and verbal, which begins with questions to gain an understanding of the child’s development and capability for an interview. A trained licensed clinical social worker conducts the interview using the RADAR interview technique (Recognizing Abuse Disclosure types and Responding; Everson et al., 2014) derived from the National Institute of Child Health and Development (NICHD) Investigative Interview Protocol (Lamb et al., 2007).

CPS and/or law enforcement involvement varies by case; either agency may or may not be involved before, during, or after an evaluation takes place. In cases in which CPS and/or law enforcement request a medical evaluation be performed at the clinic, it is common that CPS and/or law enforcement have already initiated an investigation. When a child is referred from another source, such as a community provider, law enforcement and/or CPS involvement may vary at the time the child is referred. CAN providers complied with all state reporting statutes to CPS and/or law enforcement when concerns for abuse/neglect rose to a level indicated by the statute. See below for more detailed descriptions of the possible sources of information for each factor.

#### Episodes

We defined the unit of analysis as an episode, with each episode including all notes related to one incident of suspected maltreatment for a focal child, even if these notes related to multiple healthcare encounters. Encounters for the same medical record number that occurred within 60 days of each other were assumed to be related to the same incident. Approximately 10 percent of episodes had more than one encounter. For episodes with more than one encounter, the mean number of encounters was 2.1 (*SD*=0.31). The mean number of days between the first and second encounter in an episode was 21.3 days (*SD*=20.1). Episodes were categorized as inpatient only, inpatient with outpatient clinic follow-up, and outpatient clinic only.

### Chart Abstraction Codes and Procedures

#### Coding Scheme Development

Using an interdisciplinary team-based approach to qualitative conceptual content analysis, the development of a codebook for chart abstraction was led by a pediatrician specializing in child abuse and neglect and a researcher with a Master of Social Work (MSW). The basic structure we used for organizing codebooks allowed for flexibility throughout the coding process that facilitates coder training (MacQueen et al., 1998). First, the researcher reviewed 100 clinic visit notes to develop initial coding themes with consultation from the child abuse pediatrician. Two additional coders then joined the researcher and pediatrician through an iterative process to identify codes for analysis to examine the occurrence of selected terms in the data. When they reached a point that no new themes were identified, the codebook was developed to specify coding decisions, alleviate ambiguity, and ensure inter-coder reliability.

##### Coder Training

A team of 11 coders including doctoral- and masters-level researchers (*n* = 6), medical students (*n* = 4), and a research assistant (*n* = 1) were trained. To ensure consistency, each coder completed a training set of 15 cases and compared them to the “gold standard” master set coded by the researcher with an MSW and the child abuse pediatrician. Coders were required to reach greater than 90% agreement with the master set during training and review any errors with master coders before initiating abstraction of the full dataset.

Notes were initially coded in NVivo Pro databases (QSR International Pty Ltd, 2018). These data were then extracted into a secure REDCap database (Harris et al., 2019; Harris et al., 2009), where coders flagged any question or potential coding discrepancy. Comments and questions captured within REDCap were addressed by the MSW, data analyst, or pediatrician as needed. The coding team met regularly to discuss any uncertainties, questions, and issues that arose, and they updated the codebook based on any decisions made. This process facilitated a better means for quantitative analysis and allowed for a comparison between coding within REDCap (Harris et al., 2019; Harris et al., 2009) and NVivo (QSR International Pty Ltd, 2018). In an effort to reduce any potential secondary trauma from reading reports of alleged child maltreatment, coders were asked to only code reports for a maximum of two hours per day based on recommendations for researchers to plan a workload to allow space and time in between exposure to traumatic materials (Coles et al., 2010). This restriction was also intended to promote accuracy by avoiding coding fatigue.

### Analysis of Interrater Agreement

We used Gwet’s agreement coefficient (Gwet, 2014) to analyze interrater agreement for each code. Interrater agreement is defined as “the propensity for two or more raters to independently classify a given subject into the same predefined category” (Klein, 2018). We used the -kappaetc-command in STATA 16.0 (StataCorp, 2019) to calculate this coefficient.

## Results

### Demographics

Age, sex, and race/ethnicity (Non-Hispanic Black, Non-Hispanic White, Hispanic, and Other), and health insurance type (Medicaid and others) of the focal child were coded from the child’s electronic health record. Our sample included 1,397 unique identifiable children (see Figure 1 for flow diagram). Table 1 presents the distribution of child demographic characteristics. There were more girls (61.7%) than boys (38.3%). Non-Hispanic Black children were represented at a higher proportion (57.3%) than Hispanic (21.5%) or non-Hispanic White children (13.9%). The youngest children (ages 0-3) made up 27% of the sample. Most children (70.7%) were insured by Medicaid. More girls were in the 12-17 years group (80.0%) than in the other two age groups (54.4% for 0-3-year-olds and 57.5% for 4-11-year-olds). In addition, a lower percentage of children 12-17 years old (64.0%) were enrolled in Medicaid than other age groups (78.4% for 0-3-year-olds and 69.6% for 4-11-year-olds).

**Figure 1.**
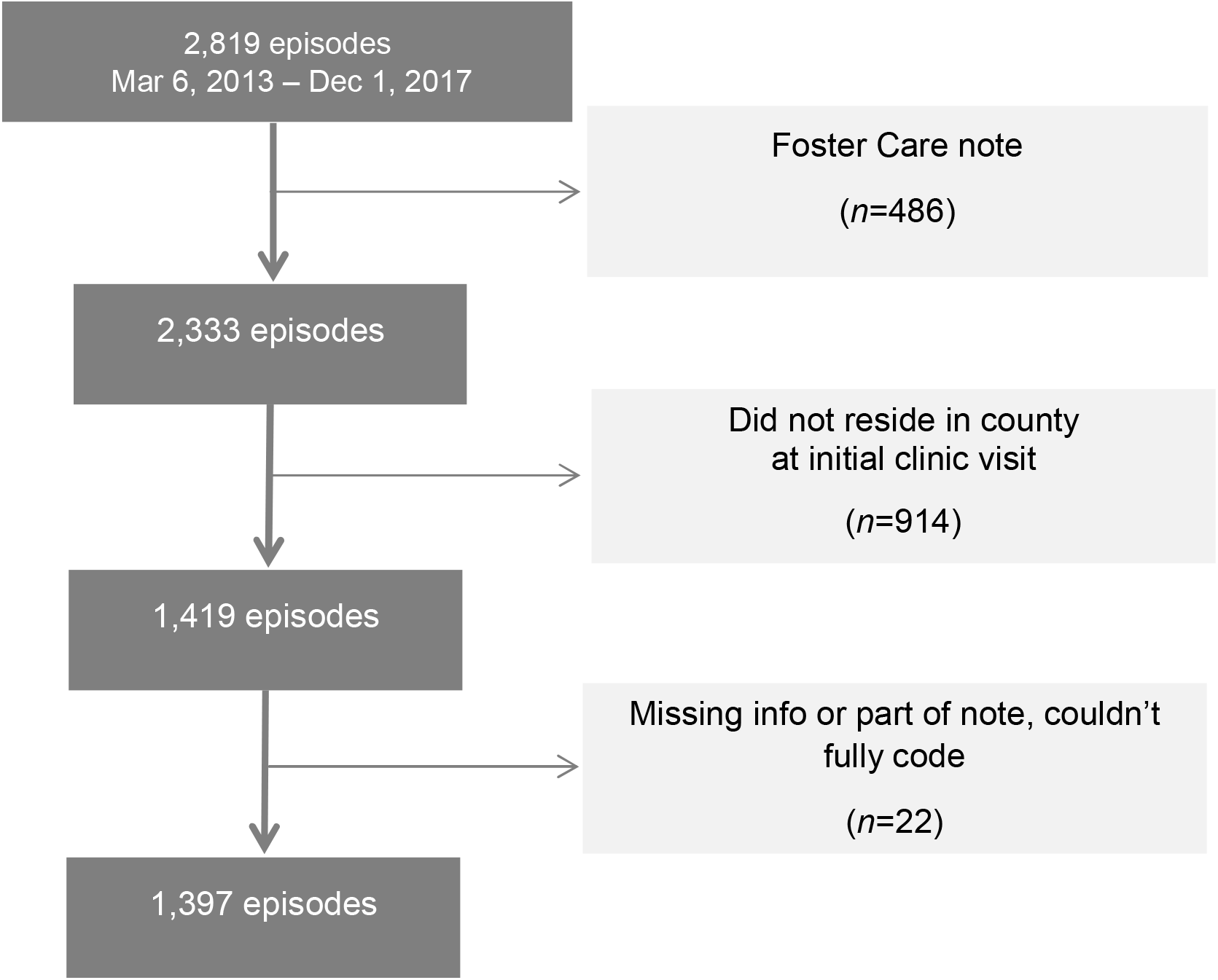
Sample Flow Diagram

**Table 1.** Sample Demographic Characteristics

| Characteristics | Child Total<br>(*N* = 1,397) | 0-3 years<br>(*n* = 375) | 4-11 years<br>(*n* = 708) | 12-17 years<br>(*n* = 314) | *p* |
| --- | --- | --- | --- | --- | --- |
| Sex |  |  |  |  | 0.000 |
| Male | 535 (38.3) | 171 (45.6) | 301 (42.5) | 63 (20.1) |  |
| Female | 862 (61.7) | 204 (54.4) | 407 (57.5) | 251 (80.0) |  |
| Race |  |  |  |  | 0.036 |
| Non-Hispanic Black | 800 (57.3) | 219 (58.4) | 394 (55.7) | 187 (59.6) |  |
| Non-Hispanic White | 194 (13.9) | 64 (17.1) | 94 (13.3) | 36 (11.5) |  |
| Hispanic | 300 (21.5) | 60 (16.0) | 171 (24.2) | 69 (22.0) |  |
| Other | 103 (7.4) | 32 (8.5) | 49 (6.9) | 22 (7.0) |  |
| Age |  |  |  |  |  |
| 0-3 | 375 (26.8) |  |  |  |  |
| 4-11 | 708 (50.7) | N/A |  |  |  |
| 12-17 | 314 (22.5) |  |  |  |  |
| Insurance type |  |  |  |  | 0.000 |
| Medicaid | 988 (70.7) | 294 (78.4) | 493 (69.6) | 201 (64.0) |  |
| All others | 409 (29.3) | 81 (21.6) | 215 (30.4) | 113 (36.0) |  |

*Note.* *P* value for $\chi^2$ tests. -, cell that is less than or equal to 10.

### Codes

The averaged Gwet’s agreement coefficients for each code ranged from 0.84 to 0.99. These coefficients indicate high coder agreement. Categories that were coded for fewer than ten cases are not shown in any table to protect identifiable data as mandated by the IRB.

#### Referral Source

The origin of the referral was coded as Child Protective Services (CPS), law enforcement, pediatrician, emergency department, other medical provider, or other. Child Protective Services (CPS) was the most frequent referral source (56.1%), followed by the emergency department (15.8%), pediatricians (12.0%), police (10.9%), hospital referrals (7.7%), and referrals by other medical professionals (2.7%) CPS referrals were the source of over 60% of cases in each of the age groups above three years. The youngest group of children (i.e., 0-3 years of age) were more likely to be referred for evaluation by the emergency department or hospitals compared with older age groups. Tables of descriptive results for all codes described here can be found in the supplementary material.

#### Family Risk Factors and Exposures

Exposures that could affect all members of a family/household were coded for each episode, including housing insecurity, domestic violence, parental criminal justice system involvement, parental substance use, parental mental health, and parental history of victimization/perpetration (see Table 2). The source of this information varies by case and depends on who accompanied the child to the clinic visit. It could include the caregiver, another family member, a foster parent, or CPS worker assigned to the case. When a CPS social worker is involved in the case, their notes may also be shared via consultation to provide a source of information. Overall, 75.9% of the families had at least one risk factor and almost 18% had four or more risk factors. Domestic violence was the most frequently reported risk factor across age groups, with almost half of all cases indicating a history (47.0%). Parental criminal involvement (38.1%) and parental incarceration (15.9%) also affected a substantial number of families. About 26.8% of parents had a reported mental health issue and 28.3% had reported a substance use problem. Parents had a history of victimization in about one quarter of cases (26.1%) and a history of perpetration in 9.5% of cases. Housing insecurity was a risk factor in 7.2% of cases, and 2.4% involved children’s exposure to a known sex offender. Parents of children 0-3 years old had higher rates of housing insecurity and mental health issues than parents of older children, while parents of children 4-11 years old had the highest percentages of domestic violence and criminal involvement among all the cases. The distribution of other risk factors was similar across age groups.

**Table 2.** Coded Risk Factors and Exposures

| Family Level Risk Factors | Code |
| --- | --- |
| Domestic violence | Captures focal child's current and historical exposure to and/or experience with violence in the household |
| Housing insecurity | Captures focal child's current and historical exposure to and/or experience with housing insecurity |
| Exposure to registered sex offender | Focal child has any exposure to a registered sex offender. Only coded in cases of suspected sexual abuse. |
| Parental incarceration | Jail, state or federal prison incarceration. This code is used when the clinician reports the person had a stay in a jail, prison, or was incarcerated. |
| Parental criminal involvement | Criminal arrests or legal charges that did not lead to incarceration |
| Parental substance use | Reports of use and misuse of substances, including alcohol. Does not include reports of possession/selling/distributing of substances. |
| Parental mental health | Reports of parent's current and historical mental health concerns. Included all factors listed below as individual level factors, as well as post-partum depression. |
| Parental history of victimization | Reports of parent experiencing maltreatment including physical, emotional, sexual abuse, or neglect. |
| Parental history of perpetration | Report of parent abusing a child physically, emotionally, or sexually. |
| Individual Level Factors | Codes |
| ADD/ADHD | Coded for presence in the notes |
| Anxiety | Includes panic attacks. Coded for presence in the notes |
| Autism Spectrum Disorder | Coded for presence in the notes for ages 3+ years |
| Bipolar Disorder | Coded for presence in the notes |
| Depression | Coded for presence in the notes |
| Developmental delay | Coded for presence in the notes for ages 3+ years |
| Oppositional Defiant Disorder | Coded for presence in the notes |
| Post-traumatic Stress Disorder (PTSD) | Coded for presence in the notes |
| Schizophrenia | Coded for presence in the notes |
| Substance use | Coded for presence of reports of use and misuse of substances, including alcohol. Does not include reports of possession/selling/distributing of substances. |
| Suicidal behavior | Acute suicidality, suicide precautions. Does not include non-suicidal self-injury. |
| Other mental health concerns | Coded for presence in the notes when clinician reports mental health concerns but does not specify the exact nature of the concern. This code may be used concurrently with specific mental health concerns. |

#### Individual Mental Health Risk Factors

Mental health disorders were based on clinical notes for the presence of more than 12 types of psychological disorders such as anxiety and depression. For the focal child, mental health diagnoses and substance use could appear as a known diagnosis found in the medical chart, or provided by another professional, or could also be a non-confirmed diagnosis reported by a caregiver. Risk factors were coded as “present” if the above definitions were met by the focal child. See Table 2 for a list of the individual factors coded for the focal child. ADD/ADHD was the most common problem reported for children (10.6%), followed by other mental health concerns (8.7%) and depression (5.9%). Around 27.0% of children had experienced at least one risk factor and 2.7% had four or more risk factors. Children 0-3 years old rarely had documented individual level risk factors, and children 12-17 years old had the highest percentage for all individual risk factors among the three age groups.

#### Child Protective Services (CPS) Involvement

Notes indicated the focal child’s current or past involvement in CPS, including foster care or kinship care. CPS involvement may be indicated by the referral source, and/or reports from CPS workers. While CAN medical providers do not have access to the CPS system or their ongoing notes, any existing CPS intake reports may be provided along with referral, and CPS workers may provide information via phone or in-person interview. Over half of the children in our cohort had an open CPS case that did not involve out-of-home placement at the time of their medical evaluation for maltreatment. About 7.3 percent and 4.8 percent of total cases involved children who were in foster care or kinship care at the time of their evaluation, respectively. Six percent of the children had a history of prior foster or kinship care. Nearly one-third of the children (31.4%) had a prior CPS history other than foster or kinship care. Compared to younger children, older children were more likely to have been in foster care or had a prior CPS history.

#### Parenting Practices

Caregivers reported on the methods they use when disciplining their children. Removal of privileges was the most frequently used parenting strategy (44.1%), followed by physical discipline (32.1%) and time out (25.4%). Physical discipline (42.9%) and time out (35.3%) were most frequently used among 4-11-year-olds, while removal of privileges (59.5%) was mostly used among the 12-17-year-olds.

#### Maltreatment Type

The CAN clinic identifies all types of child maltreatment. This includes physical abuse, neglect, medical neglect, and emotional abuse. It also encompasses maltreatment that is sexual in nature. Sex-related maltreatment can be further specified as abuse, assault, exploration, and victimization.

#### Level of Clinical Concern

The role of the CAN medical evaluation clinic is to provide a medical evaluation for children referred for concerns of abuse or neglect. Medical diagnoses, conclusions, and recommendations made for children referred to the clinic are based on the information available at the time of evaluation. Thus, the terminology used for medical conclusions and the guidelines used to determine these conclusions are based on clinical information and scientific data in the field of child abuse and neglect, and do not constitute or equate to legal terminology or conclusions regarding abuse/neglect. The diagnostic categories do not include any custody determinations, which are done by legal entities. The CAN specialty clinic performs a multidisciplinary case conference to peer review each evaluation and determine a diagnosis that describes a level of concern of maltreatment for the child. The CAN medical team, which includes child abuse pediatricians, licensed clinical social workers, and a team nurse practitioner, gathers weekly to discuss important details and determine a medical diagnosis. The goal of case review is to decrease bias and improve standardization of diagnoses. The CAN clinic team uses six levels of medical concern for cases referred for evaluation: clear and confirmed, probable, suspicious, unlikely, unknown, and no concern. Levels of concern are guidelines to promote consistency and high-quality medical evaluations. These guidelines also help inform adherence to mandatory reporting statutes and consistency of medical recommendations. Since they are guidelines, there may be variations depending on individual circumstances. Table 3 displays the diagnostic levels of clinical concern for physical abuse, neglect, and sex-related abuse with brief descriptions of each level. This table is a condensed form of a more complex document used during case review.

**Table 3.** Levels of Clinical Concern Guidelines

| Level | Description |
| --- | --- |
| Clear and confirmed | Case history includes verified medical findings of abuse/neglect. |
| Probable | Case history includes information leading to high level of medical concern for possible abuse/neglect of the child. |
| Suspicious | Case history includes information which is medically concerning for abuse/neglect. However, there are factors which limit the ability to raise the level of concern. |
| Unknown | Case history includes information which cannot rule out the possibility of abuse/neglect, and where there are factors that may leave a child at risk for abuse/neglect. |
| Unlikely | Case history includes information which leads to low concern for abuse/neglect. |
| No concern | Case history includes information that leads to no significant medical concern for abuse/neglect. |

Overall, about one third of the cases in the sample were classified as “clear and confirmed” maltreatment, and only 5.8% were classified as “unlikely or no” for maltreatment. Children 12-17 years old had the highest percentage evaluated as “clear and confirmed” (35.7%) or “probable” (29.3%), and children 0-3 years old had the highest rate for “unknown” (32.0%) or “unlikely or no” (9.6%) across age groups.

##### Physical Abuse

About 44.3% of the total sample was evaluated for physical abuse, with the youngest children (0-3 years of age) evaluated for physical abuse more frequently than children in older age groups (59.7% for 0-3 year olds, 39.4% for 4-11 year olds, and 36.9% for 12-17 year olds). Nearly 10% of all cases were classified as “clear and confirmed”, with an additional 4.8% classified as “probable”, 12.9% as “suspicious”, and 14.8% as “unknown”. Only 2.4% of cases were deemed to be “unlikely or no”. Rates of “clear and confirmed” assessments were fairly consistent across age groups but highest for 7-13-year-olds (11.5%). “Suspicious”, “unknown”, and “unlikely or no” conclusions generally decreased with increasing age of the focal child.

##### Neglect

Approximately 35.1% of our sample was evaluated for neglect. Over half of the youngest children received evaluations for neglect (51.7%), which was higher than the evaluated older age groups (30.8% for 4-11 and 35.1% for 12-17). Overall, 10.8% of all cases were assessed as “clear and confirmed” for neglect. Children 0-3 years of age had the highest rates for neglect across all the clinical levels of concern compared with older age groups.

##### Sex-related Abuse

Sexual abuse/assault/exploitation/victimization was evaluated in nearly 60% of all cases, making it the most prevalent form of abuse evaluated in our sample. The youngest children were evaluated for sex-related abuses at the lowest rates (35.2% of 0-3-year-olds), while over 70% of the 12-17-year-olds in the sample were evaluated for sex-related abuse. Overall, 8.9% of total cases were “clear and confirmed”, 19.3% were deemed “unknown”, and 4.1% “unlikely or no”. “Clear and confirmed”, “probable”, and “suspicious” cases increased with age, while “unknown” cases tended to decrease with age.

##### Medical Neglect and Emotional Abuse

Evaluation of medical neglect and emotional abuse was uncommon, with each coded in less than 4% of total cases. Only 1.3% of all cases in the entire sample were labeled “clear and confirmed” for medical neglect, and 1.6% “suspicious”. For emotional neglect, 1.3% of cases were deemed “suspicious”. All other cell sizes for medical neglect and emotional abuse were less than 10; thus, we are unable to assess age differences and did not include these findings in the supplementary information.

#### Recommendations

This code indicates whether families received recommendations for additional services from which the focal child or family members may benefit following the focal child’s CAN clinic visit. CAN medical providers may recommend multiple services for the same individual, so all recommendations were coded separately for both focal child and other individual family members. Codes in this category include therapeutic trauma-informed mental health services, CPS report, developmental evaluation, domestic violence counseling, individualized education plan (IEP), parenting classes, parenting coordinator, parent-child interaction therapy, speech assessment, substance abuse counseling, and other. See Table 4 for descriptions of each recommendation code. General mental health service was the most common service recommendation for focal children (42.9%). Therapeutic trauma-informed mental health service was the next most common recommendation (22.6%). Eight percent of children were referred for child-family evaluations, followed by “other” recommendations (8.0%), such as referral to a subspecialty provider. Adolescents 12-17 years of age were more frequently referred for therapeutic trauma-informed mental health services, CPS reports, and general mental health services than children in the younger age groups. Children 0-3 years of age were more likely to be recommended to developmental evaluation and speech assessment compared to children in older age groups.

**Table 4.** CAN Provider Recommendation Codes

| Service Recommendations | Description |
| --- | --- |
| Therapeutic trauma-informed mental health services | Specialized services provided by an expert who is certified/licensed in specific trauma therapy techniques, such as trauma-focused cognitive behavioral therapy |
| Child-Family Evaluation | Brief, specialized forensic evaluation in cases that have not been determined through the standard CPS investigation or CME. |
| Child Medical Evaluation | Recommend a medical consultation performed by a qualified medical expert rostered with the state Child Medical Evaluation Program to assist with determining the most appropriate medical diagnoses and treatment plan for a child when it is suspected that child is being abused or neglected by a caretaker. Usually recommended for siblings or other children in the home. |
| CPS report | File a report with CPS if one has not already been filed. |
| Developmental evaluation | Assessment of various aspects of the focal child's functioning, including areas such as cognition, communication, behavior, social interaction, motor and sensory abilities, and adaptive skills. |
| Domestic violence counseling | Specialized counseling/therapy from a qualified professional who has training and experience treating individuals who experience(d) intimate partner violence. |
| General mental health services | Any mental health services provided by a licensed provider for a specific age group. |
| Individual Education Program (IEP) | Coordinating with a team of individuals from various educational disciplines to create a plan to ensure that a child with an identified disability receives specialized instruction and services. |
| Parenting coordinator | A neutral third-party who helps parents involved in high-conflict custody cases make day-to-day decisions |
| Parent Child Interaction Therapy | Evidence-based behavior parent training treatment for young children with emotional and behavioral disorders. PCIT places emphasis on improving the quality of the parent-child relationship and changing parent-child interaction patterns. |
| Speech assessment | Recommend assessment to establish whether a child has any specific speech, language, or potential communication disorders. |
| Substance abuse counseling | Services provided by a qualified professional trained and experienced in providing support to patients suffering from drug or alcohol dependency and educating families in the best ways to help in the recovery process. |
| Other | Recommendations that do not have an existing recommendation code |

## Discussion

Child abuse and neglect medical providers who perform medical evaluations assess multiple factors across multiple contextual levels and conduct multi-disciplinary peer review of cases to determine the level of clinical concern maltreatment. In this study, we report the development of a qualitative coding system to extract information from child abuse and neglect medical evaluation clinic notes. Using the system developed by an interdisciplinary team to capture individual and family-level risk factors, our coders’ agreement coefficients were high overall, ranging from 0.84 to 0.99. This study demonstrates the development and implementation of the coding system and highlights the potential value of extracting variables to assess case characteristics and patterns from medical provider notes. Data collected in this manner, while effortful and time-consuming, can help guide best clinical practices and further research in child maltreatment prevention and treatment.

Assessing aggregate information coded from medical provider notes provides a useful overview of trends in cases evaluated for child abuse and neglect, as demonstrated by previously published studies that have similarly extracted information from child abuse pediatrician (CAP) case notes. Coded data have been used by researchers to identify different notetaking approaches used by CAPs (Keenan & Campbell, 2015), to provide descriptive data on the incidence of child abuse and neglect cases and subsequent interventions in the Netherlands (Teeuw et al., 2017), to investigate legal consequences of child abuse (Hendrix et al., 2020), and to describe medical consultations by CAPs in a hospital setting in the U.S. (Hicks et al., 2020; Johnson et al., 2021). While previous studies were conducted primarily by medical professionals, our interdisciplinary team included diverse perspectives from researchers in health policy, public health, social work, and developmental psychology, in addition to medical providers. Our study builds upon previous work by including a substantial number of adolescent cases, taking a developmental-contextual risk and resilience approach, and including measures of cumulative risk at the individual and family level. In addition, this is the first study, to our knowledge, that includes detailed information on the development and implementation of the coding system used to extract data from CAN medical evaluation notes. We hope that by documenting our process and the coding materials, it will enable future research teams to use these methods and reduce costs and time for conducting related research.

Like previous studies in the U.S., the expert panel of CAN medical providers in the current study rated fewer than half of total referred cases as *clear and confirmed* or *probable* for any type of maltreatment. In fact, the overall rate in our study (33%) was lower than previous studies rating confirmed abuse cases (Hendrix et al., 2020; Hicks et al., 2020; Johnson et al., 2021). This reflects variability across different medical centers in terms of percentage of cases assessed to have a low versus high likelihood of abuse, as discussed by Johnson et al. (2021). In the current study, there were very few cases assessing medical neglect or emotional abuse, which were not examined in the studies above.

To situate our findings in a developmental-contextual framework (Belsky, 1993), we presented the results by age group (0-3 year-olds, 4-11 year-olds, and 12-17 year-old adolescents). Our findings indicate that the youngest children were evaluated at higher rates than older children. They had the highest percentages for evaluation of physical maltreatment and neglect, and their notes documented several risk factors, such as housing insecurity and mental health issues, at rates higher than those observed among older children. However, the youngest children also had the highest rate for a clinical concern of “unknown” or “unlikely or no”. Children under age four are particularly vulnerable to experiencing maltreatment as they rely on caregivers to provide nearly all their needs (Conrad□Hiebner et al., 2019). Young children are unable to protect themselves and are largely unable to communicate concerns of physical injuries or neglect to an adult. Thus, young children represent a disproportionate percentage of victims and a unique population for early identification and prevention of maltreatment efforts. On the other hand, the adolescents in the 12-17 age group were evaluated most frequently for sex-related abuse and had the highest rates of clear and confirmed sex-related abuse. This mirrors research indicating that the risk of child sexual abuse increases with age, particularly in adolescence (Putnam, 2003), with a rise in victimization with each additional year from 15 to 17 (Finkelhor et al., 2014). Further, unlike young children, adolescents can vocalize concerns, and to some extent, advocate for their own needs.

Most of the individual-level risk factors coded from medical providers’ notes did not apply to the children under age four (e.g., ADHD, anxiety, depression, substance use). The highest rates of individual risk factors were among adolescents. One in five adolescents had reported diagnosis of ADHD, which is higher than the 13.6% national prevalence rate for 12-17-year-olds (Danielson et al., 2021). Depression and anxiety typically increase with age (Ghandour et al., 2019), and the prevalence of depression (20%) in our adolescent sample is much higher than that in the general population of children (4.9%) aged 6-17 years (Bitsko et al., 2018). Further, anxiety has usually been found to be more prevalent than depression (Bitsko et al., 2018; Ghandour et al., 2019), while we found contrary results. Sixteen percent of adolescents’ case notes also mentioned suicidal behavior. Overall, the 12-17-year-old group had the greatest number of individual risk factors. Future research should focus on adolescence as a particular time of mental health risks among those who have experienced maltreatment.

Family-level risk factors were less dependent on a child’s age than child-level risk factors. We found that almost half (47%) of all children had a history of domestic violence in the home. Parental history of victimization, parent mental health issues, and parent substance use were risk factors present in about a quarter of cases. The notes documented multiple family risk factors more often than individual factors. Consistent with prior studies, cases with a high concern for maltreatment often presented with family-level risk factors, including a history of experiencing domestic violence, parental mental health issues, parental history of victimization, and parental substance use (Coulter & Mercado-Crespo, 2015; Dubowitz et al., 2011; Kelley et al., 2015; Victor et al., 2018). These risk factors often co-occur in a family and may cumulatively contribute to children’s adverse outcomes (Horwitz et al., 2011; Patwardhan et al., 2017).

An understanding of the contextual information related to potential incidents of abuse or neglect is crucial when researching, diagnosing, and treating child maltreatment. It is important to note, however, that there is some evidence that inclusion of social information may influence diagnosis in child maltreatment cases. Recent qualitative research suggests that the inclusion of social information in consultation notes may influence medical diagnoses regarding abuse. Keenan et al. (2017) found that the presence of social risk factors when evaluating injuries influenced child abuse consultants’ diagnosis of abuse among poor and minority children, especially in ambiguous cases. Similarly, Olson et al. (2018) found that child abuse clinicians’ consultation notes detail a rich social history that includes many domains of the child and families’ social ecology; however, information recorded was not always related to known population risk indicators (e.g., negative impression of caregivers). In both studies the authors suggested that uniform reporting styles and tools such as checklists and peer review show promise in minimizing bias, but continued research is warranted to understand how child abuse medical providers use information on patients’ social history and circumstances to understand and document their findings.

In the current study, CAN medical providers conducted multidisciplinary case conferences for peer review on each evaluation, which should help reduce bias by standardizing the process for all patients. Examining CAN medical providers’ notes through qualitative data mining may aid the development of a best practice model for managing and documenting child abuse and neglect consultations.

In addition to its strengths, this study has limitations. First, data were from only one clinical setting. Because the process whereby medical providers take family history may differ across practice settings (Johnson et al., 2021), the available information may differ by clinician. Second, the information gathered relied on intake interviews between providers and caregivers or children, and thus depended on the quality of communication and documentation in medical consultation notes. This may be subject to respondents’ willingness to share and their awareness of the child’s exposures. Thus, the absence of documented recording of social histories (e.g., history of parental childhood victimization, use of physical discipline, etc.) does not necessarily indicate the child’s family did not experience certain adversities. Third, the timing of an issue (historical and/or ongoing) and the exact nature of how affected a child was by various adversities was unclear. For example, a parent may have substance use issues and may vary in the amount of contact with the child. Similarly, a parent may have varying levels of criminal involvement—which vary in how it affects the child’s environment. Finally, a limitation is that the guidelines used by the CAN medical providers in this study have not been validated. The guidelines are used to guide the provision of care and resources, to facilitate discussion to mitigate bias and be consistent, and as internal continuous quality improvement, but validity testing has not yet been done.

## Conclusions

This study begins to examine the reported health and social conditions of children who are referred to a child abuse and neglect clinic. By hand coding notes from these evaluations, we unveiled information that medical providers may use to inform their care of children. This information offers details that are otherwise unobservable from fields commonly used to assess meaning from EHR databases such as categorical information on encounter type or ICD diagnosis. Findings demonstrate that medical providers are gathering rich information on caregiver needs and behaviors (e.g., substance use, mental health, parenting style) and family factors (e.g., criminal involvement, housing insecurity) that are risks for maltreatment (Hunter & Flores, 2021), and which may be amenable to service provision. Interestingly, about 20 percent of the children referred to the clinic had been or were currently in foster or kinship care (not including those who were referred for their foster care evaluation). Thirty percent had a prior social service report that did not involve placement outside the home. The extent to which medical systems can work with families and community partners to address social drivers of health and risk factors for experiencing maltreatment is important, although we must take care not to conflate family needs and risk factors with child maltreatment (Keenan et al., 2017).

It is valuable and feasible to extract meaningful and substantial data from child abuse and neglect medical evaluation clinic notes, but it is time- and labor-intensive. Child abuse pediatricians and other medical providers in this area should consider upgrading their documentation to include flowsheets or data input areas that increase the ease of extracting data. Research should continue to examine how medical providers make use of this information, with a priority to reducing any biases. Framing these data within a developmental-contextual lens has potential to promote partnership between health systems, community partners, and families. In aggregate, this information might inform the underlying needs of families and better position communities and health care systems to address them.

## Data Availability

All codebook data are available upon reasonable request to the authors, but the health record data is not available due to ethical reasons.

