## Supplemental Tables for "What Do Child Abuse and Neglect Medical Evaluation Consultation Notes Tell Us?"

**Supplemental Files**

**Table A1**
*Referral Sources, CPS Involvement, and Recommendations*

| Referral Source | Total  (*N* = 1397) | 0-3 years  (*n* = 375) | 4-11 years  (*n* = 708) | 12-17 years  (*n* = 314) | *p* |
| --- | --- | --- | --- | --- | --- |
| CPS | 783 (56.1) | 156 (41.6) | 432 (61.0) | 195 (62.1) | 0.000 |
| Police | 152 (10.9) | - | 95 (13.4) | 47 (15.0) | 0.000 |
| Medical Provider |  |  |  |  |  |
| Emergency department | 220 (15.8) | 90 (24.0) | 71 (10.0) | 59 (18.8) | 0.000 |
| Pediatrician | 167 (12.0) | 46 (12.3) | 97 (13.7) | 24 (7.6) | 0.022 |
| Hospital referral source | 107 (7.7) | 86 (22.9) | 16 (2.3) | - | 0.000 |
| Other medical provider | 37 (2.7) | 11 (2.9) | 21 (3.0) | - | 0.416 |
| CPS Involvement | Total  (*N* = 1397) | 0-3 years  (*n* = 375) | 4-11 years  (*n* = 708) | 12-17 years  (*n* = 314) | *p* |
| Existing CPS case open but not currently in foster care or kinship care | 716 (51.3) | 181 (48.3) | 378 (53.4) | 157 (50.0) | 0.243 |
| Current foster care | 102 (7.3) | 27 (7.2) | 52 (7.3) | 23 (7.3) | 0.996 |
| Current kinship care | 67 (4.8) | 23 (6.1) | 26 (3.7) | 18 (5.7) | 0.133 |
| Previous foster care | 46 (3.3) | - | 22 (3.1) | 21 (6.7) | 0.000 |
| Previous kinship care | 41 (2.9) | - | 21 (3.0) | 14 (4.5) | 0.086 |
| Prior CPS history other than foster care or kinship care | 439 (31.4) | 74 (19.7) | 249 (35.2) | 116 (36.9) | 0.000 |
| Services Recommendations | Total  (*N* = 1397) | 0-3 years  (*n* = 375) | 4-11 years  (*n* = 708) | 12-17 years  (*n* = 314) | *p* |
| Therapeutic trauma-informed mental health services | 315 (22.6) | 28 (7.5) | 190 (26.8) | 97 (30.9) | 0.000 |
| Child-Family Evaluation | 113 (8.1) | - | 76 (10.7) | 30 (9.6) | 0.000 |
| Child Medical Evaluation | 24 (1.7) | - | - | - | 0.023 |
| CPS report | 18 (1.3) | - | - | 12 (3.8) | 0.000 |
| Developmental evaluation | 50 (3.6) | 20 (5.3) | 25 (3.5) | - | 0.031 |
| General mental health services | 599 (42.9) | 41 (10.9) | 371 (52.4) | 187 (59.6) | 0.000 |
| Parenting coordinator | 13 (0.9) | - | - | - | 0.091 |
| Parent Child Interaction Therapy | 18 (1.3) | - | 12 (1.7) | - | 0.070 |
| Speech assessment | 44 (3.2) | 16 (4.3) | 27 (3.8) | - | 0.004 |
| Other | 111 (8.0) | 17 (4.5) | 70 (9.9) | 24 (7.6) | 0.008 |

*Note.* P value for χ^2^ tests. -, cell that is less than or equal to 10. CPS, Child Protective Services; these cells are not mutually exclusive. Due to small sample size, the results for Carolina Outreach, Care Coordination for Children, Children’s Developmental Services Agencies, domestic violence counseling, Individualized Education Program, parenting classes, and substance abuse counseling are not shown.

**Table A2**
*Individual Level Risk Factors and Exposures*

| Risk factors | Child Total  (*N* = 1397) | 0-3 years  (*n* = 375) | 4-11 years  (*n* = 708) | 12-17 years  (*n* = 314) | *p* |
| --- | --- | --- | --- | --- | --- |
| ADD/ADHD | 148 (10.6) | - | 86 (12.2) | 62 (19.8) | 0.000 |
| Anxiety | 37 (2.7) | - | 17 (2.4) | 20 (6.4) | 0.000 |
| Autism Spectrum Disorder | 24 (1.9) | - | 19 (2.7) | - | 0.026 |
| Bipolar Disorder | 13 (0.9) | - | - | - | 0.000 |
| Depression | 82 (5.9) | - | 20 (2.8) | 62 (19.8) | 0.000 |
| Developmental delay | 69 (5.4) | - | 40 (5.7) | 20 (6.4) | 0.349 |
| Oppositional Defiant Disorder | 41 (2.9) | - | 20 (2.8) | 21 (6.7) | 0.000 |
| Positive substance use | 40 (2.9) | - | - | 39 (12.4) | 0.000 |
| Postpartum depression | - |  |  |  |  |
| PTSD | 24 (1.7) | - | - | 13 (4.1) | 0.000 |
| Schizophrenia | - | - | - | - | 0.038 |
| Suicidal behavior | 61 (4.4) | - | 12 (1.7) | 49 (15.6) | 0.000 |
| Other mental health concerns | 122 (8.7) | 11 (2.9) | 38 (5.4) | 73 (23.3) | 0.000 |
| Number of risk factors, *m* (*sd*) | 0.5 (1.0) | 0.1 (0.2) | 0.4 (0.8) | 1.3 (1.6) | 0.000 |
| Number of risk factors |  |  |  |  | 0.000 |
| 0 | 1020 (73.0) | 355 (94.7) | 534 (75.4) | 131 (41.7) |  |
| 1 | 209 (15.0) | 19 (5.1) | 112 (15.8) | 78 (24.8) |  |
| 2 | 81 (5.8) | - | 33 (4.7) | 47 (15.0) |  |
| 3 | 49 (3.5) | - | 19 (2.7) | 30 (9.6) |  |
| 4 and more | 38 (2.7) | - | - | 28 (8.9) |  |
| Any risk factor | 377 (27.0) | 20 (5.3) | 174 (24.6) | 183 (58.3) | 0.000 |

*Note.* *P* value for χ^2^ tests. -, cell that is less than or equal to 10. *m*, mean. *sd*, standard deviation. Due to small sample size, the results for anger management, anti-social personality disorder, borderline personality disorder, conduct disorder, homicidal tendencies, impulse control disorder, low IQ, and reactive attachment disorder are not shown.

**Table A3**
*Family Level Risk Factors and Exposures*

| Risk factors | Total  (*N* = 1397) | 0-3 years  (*n* = 375) | 4-11 years  (*n* = 708) | 12-17 years  (*n* = 314) | *p* |
| --- | --- | --- | --- | --- | --- |
| Domestic violence | 657 (47.0) | 162 (43.2) | 362 (51.1) | 133 (42.4) | 0.008 |
| Housing insecurity | 101 (7.2) | 37 (9.9) | 51 (7.2) | 13 (4.1) | 0.015 |
| Exposure to registered sex offender | 30 (2.4) | - | 21 (3.0) | - | 0.286 |
| Parental incarceration | 222 (15.9) | 62 (16.5) | 112 (15.8) | 48 (15.3) | 0.903 |
| Parental criminal involvement | 532 (38.1) | 149 (39.7) | 289 (40.8) | 94 (29.9) | 0.003 |
| Parental mental health | 375 (26.8) | 129 (34.4) | 179 (25.3) | 67 (21.3) | 0.000 |
| Parental substance use | 395 (28.3) | 119 (31.7) | 201 (28.4) | 75 (23.9) | 0.074 |
| Parental history of victimization | 364 (26.1) | 84 (22.4) | 203 (28.7) | 77 (24.5) | 0.064 |
| Mother history of victimization | 333 (23.8) | 74 (19.7) | 187 (26.4) | 72 (22.9) | 0.045 |
| Father history of victimization | 47 (3.4) | 16 (4.3) | 25 (3.5) | - | 0.219 |
| Parental history of perpetration | 132 (9.5) | 25 (6.7) | 73 (10.3) | 34 (10.8) | 0.095 |
| Mother history of perpetration | 37 (2.7) | - | 17 (2.4) | - | 0.772 |
| Father history of perpetration | 99 (7.1) | 16 (4.3) | 58 (8.2) | 25 (8.0) | 0.045 |
| Number of risk factors,  *m (sd)* | 2.0 (1.7) | 2.0 (1.8) | 2.1 (1.6) | 1.7 (1.5) | 0.004 |
| Number of risk factors |  |  |  |  | 0.000 |
| 0 | 308 (22.1) | 70 (18.7) | 151 (21.3) | 87 (27.7) |  |
| 1 | 243 (17.4) | 42 (11.2) | 131 (18.5) | 70 (22.3) |  |
| 2 | 238 (17.0) | 35 (9.3) | 143 (20.2) | 60 (19.1) |  |
| 3 | 233 (16.7) | 41 (10.9) | 139 (19.6) | 53 (16.9) |  |
| 4 and more | 246 (17.6) | 58 (15.5) | 144 (20.3) | 44 (14.0) |  |
| Any risk factor | 1060 (75.9) | 276 (73.6) | 557 (78.7) | 227 (72.3) | 0.043 |

*Note.* *P* value for χ^2^ tests. -, cell that is less than or equal to 10. *m*, mean. *sd*, standard deviation.

**Table A4**

*Parenting Practice by Age*

| Parenting practice | Total  (*N* = 1397) | 0-3 years  (*n* = 375) | 4-11 years  (*n* = 708) | 12-17 years  (*n* = 314) | *p* |
| --- | --- | --- | --- | --- | --- |
| Physical discipline | 449 (32.1) | 49 (13.1) | 304 (42.9) | 96 (30.6) | 0.000 |
| Removal of privileges | 616 (44.1) | 37 (9.9) | 391 (55.2) | 188 (59.9) | 0.000 |
| Time out | 355 (25.4) | 56 (14.9) | 250 (35.3) | 49 (15.6) | 0.000 |

*Note.* *P* value for χ^2^ tests.

**Table A5**
*Physical, Neglect, and Sexual Maltreatment by Age*

| Highest Level of Clinical Concern | Total  (*N* = 1397) | 0-3 years  (*n* = 375) | 4-11 years  (*n* = 708) | 12-17 years  (*n* = 314) | *p* |
| --- | --- | --- | --- | --- | --- |
| Clear and confirmed | 383 (27.4) | 81 (21.6) | 190 (26.8) | 112 (35.7) | 0.000 |
| Probable | 209 (15.0) | 33 (8.8) | 84 (11.9) | 92 (29.3) |  |
| Suspicious | 424 (30.4) | 105 (28.0) | 239 (33.8) | 80 (25.5) |  |
| Unknown | 300 (21.5) | 120 (32.0) | 154 (21.8) | 26 (8.3) |  |
| Unlikely or no | 81 (5.8) | 36 (9.6) | 41 (5.8) | - |  |
| Physical abuse | Total  (*N* = 1397) | 0-3 years  (*n* = 375) | 4-11 years  (*n* = 708) | 12-17 years  (*n* = 314) | *p* |
| Clear and confirmed | 133 (9.5) | 24 (6.4) | 73 (10.3) | 36 (11.5) | 0.000 |
| Probable | 67 (4.8) | 24 (6.4) | 19 (2.7) | 43 (7.6) |  |
| Suspicious | 180 (12.9) | 60 (16.0) | 84 (11.9) | 36 (11.5) |  |
| Unknown | 206 (14.8) | 90 (24.0) | 98 (13.8) | 18 (5.7) |  |
| Unlikely or no | 33 (2.4) | 26 (6.9) | - | - |  |
| Not evaluated | 778 (55.7) | 151 (40.3) | 429 (60.6) | 198 (63.1) |  |
| Neglect | Total  (*N* = 1397) | 0-3 years  (*n* = 375) | 4-11 years  (*n* = 708) | 12-17 years  (*n* = 314) | *p* |
| Clear and confirmed | 151 (10.8) | 60 (16.0) | 63 (8.9) | 28 (8.9) | 0.000 |
| Probable | 59 (4.2) | 19 (5.1) | 25 (3.5) | 15 (4.8) |  |
| Suspicious | 206 (14.8) | 67 (17.9) | 109 (15.4) | 30 (9.6) |  |
| Unknown | 53 (3.8) | 29 (7.7) | 19 (2.7) | - |  |
| Unlikely or no | 22 (1.6) | 19 (5.1) | - | - |  |
| Not evaluated | 906 (64.9) | 181 (48.3) | 490 (69.2) | 235 (64.9) |  |
| Sexual Abuse Related | Total  (*N* = 1397) | 0-3 years  (*n* = 375) | 4-11 years  (*n* = 708) | 12-17 years  (*n* = 314) | *p* |
| Clear and confirmed | 124 (8.9) | - | 64 (9.0) | 58 (18.5) | 0.000 |
| Probable | 127 (9.1) | - | 55 (7.8) | 68 (21.7) |  |
| Suspicious | 243 (17.4) | 31 (8.3) | 145 (20.5) | 67 (21.3) |  |
| Unknown | 269 (19.3) | 81 (21.6) | 160 (22.6) | 28 (8.9) |  |
| Unlikely or no | 57 (4.1) | 14 (3.7) | 40 (5.7) | - |  |
| Not evaluated | 577 (41.3) | 243 (64.8) | 244 (34.5) | 90 (28.7) |  |

*Note.* Medical neglect and emotional neglect not shown due to small sample sizes. Children may be evaluated for multiple types of abuse and neglect during a single episode. *P* value for χ^2^ tests. -, cell that is less than or equal to 10.
